# Coxsackievirus A24: A Causative Agent of Acute Haemorrhagic Conjunctivitis Outbreak in Dar es Salaam, Tanzania, between January and February 2024

**DOI:** 10.1101/2025.04.28.24315795

**Authors:** Lawrence A. Mapunda, Peter van Heusden, Robert Baluhya, Olympia G. Machange, Ambele Mwafulango, Monica Fredrick Francis, Aziz Ituka, Omega Machange, Adela Kisanga, Edna E. Mgimba, Hamza H. Matimba, Dennis M. Kado, Elson E. Kimaro, Ramadhani A. Libenanga, Jackson Peter Mushumbusi, Hamisi M. Swalehe, Ibrahim Mauki, James Hellar, Shimba Henerico, Maria E. Kelly, Nyambura Moremi

**Author notes:** **Corresponding Author** Lawrence Amon Mapunda. These authors contributed equally to this work and share first authorship (Lawrence A. Mapunda, Peter van Heusden).

## Abstract

**Background:** On 15 January 2024, the Ministry of Health (MOH) reported a notable outbreak of acute viral conjunctivitis in the city of Dar es Salaam. In this study we investigated the causative agent of acute conjunctivitis outbreak which occurred in Dar es Salaam between Jan 2024 and Feb 2024.

**Methodology:** We conveniently collected a pair of eye swab samples from twenty-five patients presenting with symptoms of acute conjunctivitis at clinics in Dar es Salaam during the outbreak. One for bacterial culture and another for molecular detection. MacConkey and Blood agar were used to culture patient samples to identify possible bacterial causes. A Multiplex Real-time RT-PCR targeting Adenovirus, Metapneumovirus, Human Enterovirus and Parainfluenza. Positive samples with Ct value less than 30 for enterovirus were subjected to genomic sequencing by Illumina Miseq after library preparation and enrichment by Illumina’s Respiratory Pathogen ID/AMR enrichment panel kit as per manufacturer’s instructions (RPIP kit), followed by bioinformatics and phylogenetic analysis.

**Results:** Out of 25 samples, nine samples were positive for human enterovirus by Real-time PCR. Of the nine positive samples four were sequenced and Coxsackievirus A24 variant, was identified as the most probable cause of the outbreak. Phylogenetic analysis showed that samples collected during the outbreak (28AO and 26HB) belong to a clade with Coxsackievirus A24 sequences from France Mayotte from a 2024 outbreak, also close to sequences from neighbouring East African countries (Uganda, Kenya) as well as South American countries (Brazil, Mexico, and French Guiana).

**Conclusion:** This study demonstrated the utility of genomic epidemiology in identifying Coxsackievirus A24 as the pathogen responsible for the outbreak and its circulation within the East African region.

## Introduction

Most cases of acute conjunctivitis globally are caused by viruses(1), with Epidemic keratoconjunctivitis (EKC) and Pharyngoconjunctival fever (PCF) caused by adenovirus (2,3) while Acute haemorrhagic conjunctivitis (AHC) is most commonly caused by enteroviruses, Coxsackievirus A24 and Enterovirus 70 (4–6). Herpetic conjunctivitis is caused by herpes simplex virus (HSV) (7).

Coxsackievirus A24 and Enterovirus 70 are picornaviridae family enteroviruses, single-stranded non-enveloped RNA viruses which cause Acute haemorrhagic conjunctivitis. Other enterovirus genus viruses cause a range of symptoms from fever, rash, mild respiratory symptoms like cough, sore throat, runny nose, to mild gastrointestinal symptoms such as nausea, vomiting and diarrhoea (8). Enteroviruses are also known to cause more severe conditions like polio (Enterovirus C) (9), myocarditis (Coxsackievirus B)(10) and aseptic meningitis (Echovirus) (11).

Acute haemorrhagic conjunctivitis (AHC) is an extremely contagious type of viral conjunctivitis. It is commonly associated with large epidemics worldwide, especially in the tropical and subtropical regions, in humid weather and in densely populated areas (12,13). Although AHC infections are self-limiting, they usually have adverse effects on the education sector, workforce, and economic activities (12). In addition, surge of AHC cases in a Healthcare facility usually leads to straining of facilities’ resources, impeding capacity of the health facility to deliver other essential health services during an outbreak.

Patients with AHC often present with rapid onset of eye pain, foreign body sensation, swollen eyelids, subconjunctival haemorrhage, excessive tearing and sensitivity to light, symptoms which usually resolve in a period of one to two weeks (4,14). Fluids from an infected person’s eyes are highly contagious. AHC infections spread from person to person by hand-to eye-to-hand contact and infected fomite.

Acute haemorrhagic conjunctivitis was first reported in Ghana in 1969 (15). From there it spread to other parts of Africa and South-Eastern Asia, India and globally. In Western hemisphere, the epidemic of AHC occurred first in 1981 (4). Outbreaks of conjunctivitis continue to occur globally with recent major outbreaks reported in 15 of the 21 Global Burden of Disease regions. Despite that, outbreaks of AHC are commonly reported without information about etiologic pathogens responsible for conjunctivitis outbreak and are typically not identified (16).

On January 15, 2024, Tanzania Ministry of Health communicated a primary investigation report of an outbreak of viral conjunctivitis, also known as red eye disease which was reported in the city of Dar es Salaam in late December 2023 prompting an investigation. The Ministry of Health declared an outbreak of acute viral conjunctivitis with confirmed patients presenting with the symptoms of red, itchy, and burning eyes, as well as swelling, sensitivity to light and the presence of white or yellow discharge. Some individuals also experienced more symptoms such as severe headaches, excessive fatigue and swollen lymph nodes in the head and neck. The report stated that, between Dec. 22, 2023, and Jan. 11, 2024, the number of patients clinically confirmed to have red eye disease rose sharply from previously reported 17 to 869, in Dar es Salaam alone, underscoring rapid spread of the virus (14).

By January 26, 2024, the outbreak had already spread to 17 out of 30 administrative regions in Tanzania, with the reported number of cases rapidly increasing to 5,359 from 1,109 within one week. Reported cases were from Singida, Katavi, Kilimanjaro, Mara, Iringa, Njombe, Ruvuma, Simiyu, Mtwara, Lindi, Songwe, Rukwa, Mwanza, Dar es Salaam, Morogoro, Dodoma, and Pwani Regions (17). Since the data only captured symptomatic individuals who sought medical care at hospitals and clinics, the Ministry of Health suggested that the true number of cases in the community was likely higher. The Ministry of Health further advised that people in Dar es Salaam and wider Tanzania should take more precautions by implementing frequent hand washing practices to minimize the risks of transmission and infection as well as refraining from touching or rubbing eyes (17).

Conjunctivitis outbreaks have plagued Dar es Salaam since the early 1980s (18). Notably, Dar es Salaam experienced a significant viral conjunctivitis outbreak in 2010 (18). However, due to limited diagnostic abilities, etiological agents responsible for the previous outbreaks were not conclusively identified and confirmed. In this study we investigated the causative agent of acute conjunctivitis outbreak which occurred in Dar es Salaam between January and February 2024.

## Methods

### Isolate sampling

We conveniently collected a pair of eye swab samples from each of the 25 patients presenting with symptoms of acute conjunctivitis at clinics in Dar Es Salaam region. The samples were collected between 5th and 8th February 2024. One swab for virological analysis and another for bacteriological analysis were collected and transported in viral transport medium and Stuart transport medium respectively, both were sent to the National Public Health Laboratory in cold chain.

### Laboratory characterisation

#### Bacterial growth

The samples were cultured in MacConkey and blood agar following established standard microbiological techniques.

#### Molecular Detection

The samples were then subjected to RNA extraction using QIAamp Qiagen mini-RNA kit (Qiagen, German). We then performed a quantitative reverse transcriptase Polymerase Chain reaction (real time RT-PCR) using Seegene’s Allplex respiratory panel 2 (Segeene, South Korea) which targets Adenovirus, Metapneumovirus, Human Enterovirus and Parainfluenza virus.

### Sequencing and sequence assembly

Four human enterovirus positive samples with cycle threshold less than 30 were subjected to genomic sequencing using Illumina Miseq (Illumina), the sequencing library was prepared using the Illumina’s Respiratory Pathogen ID/AMR enrichment panel kit as per manufacturer’s instructions (RPIP kit). FASTQ reads were uploaded to the usegalaxy.eu Galaxy server (19) and quality trimmed using Fastp (20). Host reads were removed by mapping to the Hg38 Homo sapiens genome assembly as previously described (21). Analysis with Kraken2 (22) with the Standard-Full database (dated 2022-06-07) confirmed the presence of reads matching Enterovirus C and D in 3 out of 4 samples (28A0, 26HB and 25AM). These three samples were assembled with rnaviralSPAdes (23). The fourth sample (27YS) showed presence of *Streptococcus pneumoniae*, a known cause of bacterial conjunctivitis (24).

Full length genomes of Enterovirus C and D were downloaded from NCBI Virus (National Library of Medicine (US), National Centre for Biotechnology Information, 2024) (1,982 genomes) and searched using BLAST (25,26) using the assembled contigs from the three viral samples as queries. Two of the three samples (28A0 and 26HB) yielded high quality hits against a Coxsackievirus A24 genome, PP548240, a genome sampled in the French overseas Department of Mayotte and assembled from Oxford Nanopore sequencing.

The quality trimmed reads from samples 28A0 and 26HB were aligned against MG880745.2 (a genome sampled in Mexico and assembled using Sanger sequencing) and variants were called using snippy (27). Consensus genomes were constructed by inserting the variants called by snippy into the MG880745.2 reference genome and masking regions with low coverage with Ns using mosdepth and bcftools (28,29).

Typing with the RIVM Enterovirus typing tool (30) identified both consensus sequences as belonging to the A24v variant of Coxsackievirus A24 with the VP1 region assigned to genotype IV. Based on this classification, 69 full length genome sequences of Coxsackievirus A24 where both country and date of isolation were available were downloaded from NCBI Virus and classified using the RIVM Enterovirus typing tool. Viruses not classified as A24v (EF026081 and KU183495) were excluded from analysis, yielding a set of 67 full length genome sequences. This set of genome sequences were combined with the two consensus genomes and a multiple sequence alignment was computed using MAFFT (31). D90457.1, the prototype strain of A24v (24/70) collected in Singapore in 1970, was selected as an outgroup and a maximum likelihood tree was constructed using IQ-TREE (32) with the GTR+F+I+G4 model and 1000 bootstrap replicates. Tips and ancestral nodes were dated using the LSD2 method (33) built-into IQ-TREE2 and collected date information for all sequences in the analysis.

As many more partial Coxsackievirus A24 genome sequences are available than full length genome sequences, 1967 partial genome sequences were downloaded from NCBI Virus. The EF026081 full length sequence of the 1952 Coxsackievirus A24 Joseph strain was used to build a model for VADR (34) and this was used to annotate all full length and partial Coxsackievirus A24 sequences. The VP1 sequence was extracted from both full length and partial sequences based on VADR annotation. Partial genome sequences were selected for inclusion based on availability of date and country metadata, date greater than 1988 (for partial genome sequences), VP1 length greater than 900 base pairs and assignment to the C24v strain by the RIVM Enterovirus typing tool. This yielded a collection of 309 sequences (including the two newly generated consensus sequences). A multiple sequence alignment for all the VP1 sequences was computed using MAFFT and a dated tree constructed using IQ-TREE as above.

A comparison between the tree generated from the full genome sequences and that generated from VP1 regions showed that the full genome sequence tree was a subset of the VP1 tree and did not yield different phylogenetic results.

## Results

### Demographic characteristics of Cases

A total of 25 cases were investigated, all from Dar es Salaam, the mean age of the cases was 30, majority (80%) were males, 5 patient samples came from facilities located in Ilala district, 4 from Kigamboni district, 9 from Temeke district, and 7 from Ubungo district.

### Bacterial culture and PCR results

In the bacterial culture 17 (68%) samples had no growth, while 7 (28%) samples had *Staphylococcus epidermis* which is a normal flora, and 1 sample was *Bacillus cereus* (4%). Out of 25 samples, 9 (36%) samples were positive for human enterovirus by Real-time PCR.

### Sequencing Results

Two consensus sequences were generated (28AO and 26HB) with 84% and 64% of the 7436 base pair reference genomes (MG880745.2) covered at depth of 10 or more bases (bases with less than 10x coverage were marked as N). The samples differed from the reference genome by 320 and 246 SNPs, respectively. No insertions or deletions were detected.

The phylogenetic tree built from the VP1 samples showed that the samples collected in the outbreak (28AO and 26HB) belong to a clade (labelled as clade A in Figure 2 and shown in detail) including sequences from several African countries (Uganda, Kenya, the French overseas Department of Mayotte and Tanzania) as well as South American countries (Brazil, Mexico and French Guiana). This clade is distinct from another clade of virus circulating in several Asian countries (India, China, Thailand and others, as marked as clade B in Figure 2).

**Figure 2.**
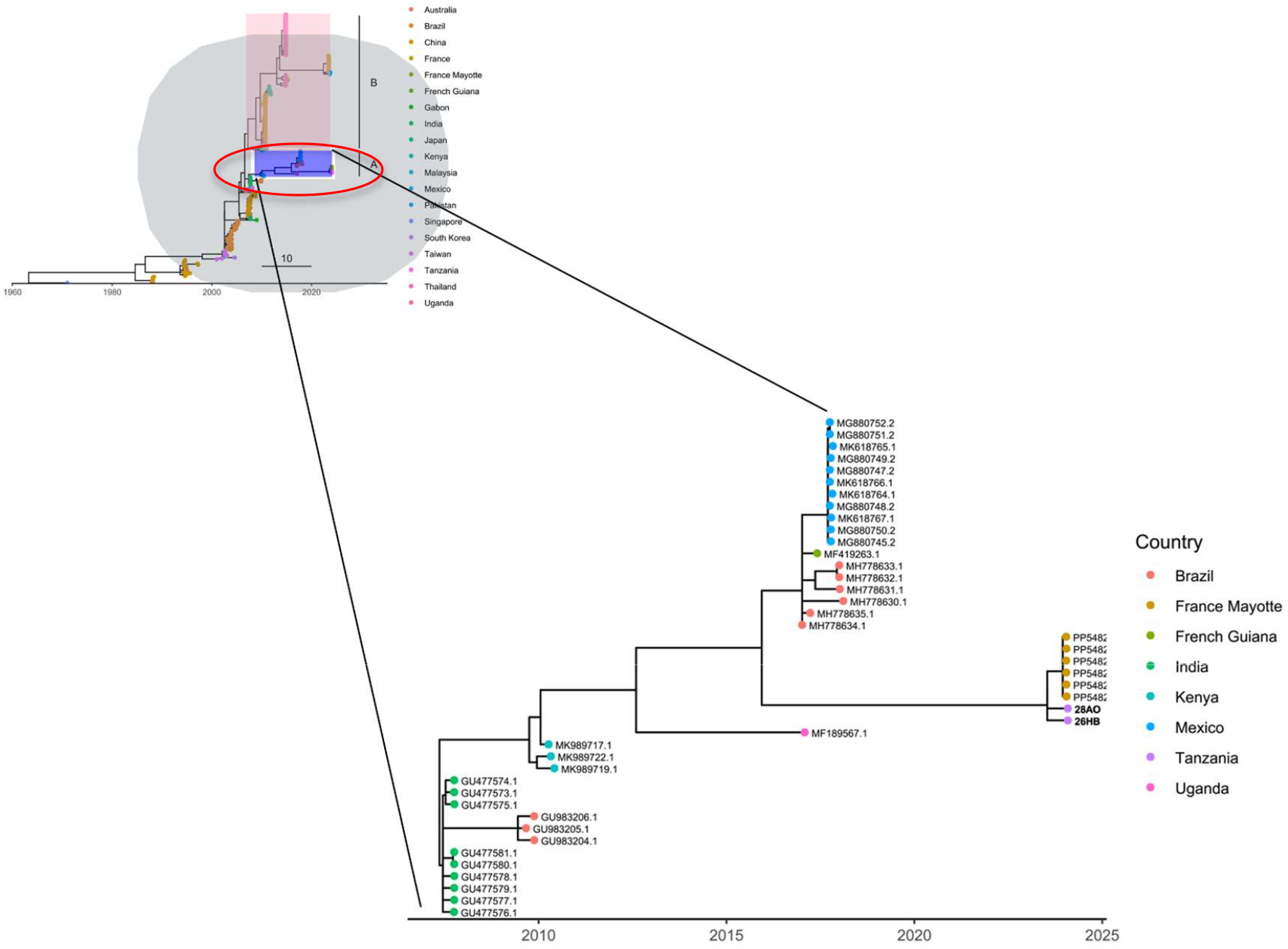
Phylogenetic tree built from VP1 sequences of Coxsackievirus A24v

## Discussion

Coxsackievirus A24(Coxsackievirus A24v) is a known cause of AHC and was first identified in Singapore in 1970 and subsequently responsible for outbreaks in Southeast Asia (35) before spreading globally including to Africa (36). In our Laboratory investigation of pair of eye swab samples collected from each of 25 patients who presented with acute haemorrhagic conjunctivitis (AHC) during the outbreak revealed that enterovirus was the most likely cause of the outbreak. Sequencing and bioinformatic analysis identified the virus as Coxsackievirus A24 variant.

Of the 25 samples, 9 (36%) tested positive for human enterovirus. These findings are consistent with the study by the Centres for Disease Control and Prevention (CDC) in 2010 which investigated a red eye syndrome outbreak in Uganda and South Sudan. In Uganda, 14 of 29 samples (48%) tested positive for enterovirus, while in South Sudan, 3 of 6 samples (50%) were positive. The same study indicated that 52% of Ugandan samples were negative for enterovirus (37), similar to the 64% negative percentage observed in our investigation. It’s worth noting that laboratory-negative results are common in viral conjunctivitis outbreaks and can be attributed to various factors, including the timing of specimen collection.

Red eye conjunctivitis outbreaks have been associated with Water Sanitation and Hygiene (WASH) practices (12,38). Conjunctivitis outbreaks can happen in favorable circumstances like hot, humid weather and crowded areas through contaminated materials and water (38). Significant flooding occurred in a number of Tanzanian regions between November and December 2023, including Arusha, Kigoma, Kagera, the Coast Region, Zanzibar, and most notably Dar es Salaam. Dar es Salaam, a significant commercial centre in East Africa is faced with overcrowding, inadequate sanitation facilities which are overwhelmed during rainy seasons and inhabitants frequently open sewage systems leading to polluting environment and runoff water (39). These factors raise the risk of diseases transmitted by faecal contamination of hands, fomite and water and pose a serious public health challenges for the city and its population. A study in Taiwan (38) reported a significant increase in incidence of diseases of the eye, skin and Gastrointestinal tract mainly due to exposure to contaminated water and materials during the flood and in the post-flood clean-up process.

Samples collected in this outbreak fit into a clade that includes samples collected in South America (Brazil, Mexico and French Guiana) as well as East Africa and surrounding islands (Kenya in 2010, Uganda in 2017 and Mayotte in 2024). While overall the phylogeny built from recent Coxsackievirus A24v VP1 sequences displays an overall ladder-like evolution, the clade is distinct from another clade including samples collected over the past decade in China, the Philippines, Thailand and Australia. This suggests that the causative agent of the recent outbreak of acute haemorrhagic conjunctivitis was a virus that has been circulating in the East Africa region for some time.

The limitation in our study was a smaller sample size and only one of seventeen regions was sampled despite the fact they all were experiencing this outbreak at the same time, however we were able to demonstrate what was the aetiological agent responsible for viral conjunctivitis in Dar es Salaam in January 2024.

## Conclusion

This study demonstrated the utility of genomics in identifying Coxsackievirus A24 as the pathogen responsible for the acute viral conjunctivitis outbreak within Dar es Salaam region. To our knowledge this is the first complete sequencing of a Coxsackievirus A24 genome by an African public health laboratory since many of the existing public genome sequences of this virus are fragmentary (less than 400bp, sometimes not including any of the VP1 region) and despite the fact that red eye disease caused by Coxsackievirus A24 is largely self-limiting, surveillance of outbreaks of unknown aetiology and of enteroviruses particular should be continued by African public health laboratories.

## Acknowledgments

The authors acknowledge the support of the Ministry of Health Tanzania for funding and capacitating the National Public Health to be able to conduct advanced molecular testing. We also would like to express our appreciation to regional eye care coordinator for Dar es Salaam, Anne Kisoka, and the technical team; Goodluck Mwanga, Kimea Myephu, Owen Mundingile, Reuben Abednego and Mussa Kichuri for the efforts they put to ensure that specimens were collected and transported to NPHL for this surveillance activity. The authors also acknowledge the value of the usegalaxy.eu service and the support of the Freiburg Galaxy Team, University of Freiburg (Germany) funded by the German Federal Ministry of Education and Research BMBF grant 031 A538A de.NBI-RBC and the Ministry of Science, Research and the Arts Baden-Württemberg (MWK) within the framework of LIBIS/de.NBI Freiburg.

## Data availability

The raw sequencing data generated in this study have been deposited in the NCBI Sequence Read Archive (SRA) under BioProject accession number PRJNA1155894.

## Ethics clearance

This study will comply with the principles stated in the Declaration of Helsinki (Ninth Revision, October 2013) and relevant national and regulatory guidelines. The study protocol was submitted to the National Health Research Ethics Committee (NatHREC) for review and approval and was granted certificate NIMR/HQ/R.8a/Vol.IX/4916. Authorization to conduct the study was obtained from the National Public Health Laboratory management.

## Author contributions

LAM and PVH: Conceived and designed the analysis, performed the analysis. LAM, PVH, RB, OGM: Wrote the paper. HS reviewed the first draft, AM, MF, AI, OM, AK, EEM, HHM, DMK, EEK, RAL, JPM, KMS, IM, JH, MEK participated in sample collection, performed laboratory testing and compiled epi data, AM, NM, supervised field and laboratory work. All authors contributed to the writing, revisions and approval of the final version of the manuscript.

## Notes

### Competing Interest Statement

The authors have declared no competing interest.

### Funding Statement

This study did not receive any funding

### Author Declarations

the study was granted ethical clearance by the National Health Research Ethics Committee (NatHREC), and was given certificate number NIMR/HQ/R.8a/Vol.IX/4916.

